# On the estimation of beat-to-beat time domain heart rate variability indices from smoothed heart rate time series

**DOI:** 10.1101/2023.10.27.23297692

**Authors:** Miguel A. Garcia-Gonzalez, Mahtab Mohammadpoor-Faskhodi, Mireya Fernandez-Chimeno, Juan J. Ramos-Castro

**Affiliations:** Department of Electronic Engineering, Universitat Politècnica de Catalunya, Campus Nord, Edifici C-4, 08034, Barcelona, Spain

## Abstract

This study tests the feasibility of estimating some time-domain heart rate variability indices (the standard deviation of the RR time series, SDNN, and the standard deviation of the differentiated RR time series, or RMSSD) from smoothed and rounded to the nearest beat per minute heart period time series using shallow neural networks. These time series are often stored in wearable devices instead of the beat-to-beat RR time series. Because the algorithm for obtaining the recorded mean heart rate in wearable devices is often not disclosed, this study test different hypothetic sampling strategies and smoothers. Sixteen features extracted from 5 minute smoothed heart period time series were employed to train, validate, and test shallow neural networks in order to provide estimates of the SDNN and RMSSD indices from freely available public databases RR time series. The results show that, using the proposed features, the median relative error (averaged for each database) in the SDNN ranges from 2% to 14% depending on the smoothness, sampling strategy, and database. The RMSSD is harder to estimate, and its median relative error ranges from 6% to 32%. The proposed methodology can be easily extended to other averaged heart rate time series, HRV indices and supervised learning algorithms

## Introduction

Heart rate variability (HRV) helps to assess the status of the autonomic nervous system (ANS) [1] and has been used for the last decades as a tool to quantify risk in a wide variety of both cardiac and non-cardiac disorders [2]. HRV reflects physiological variation in the duration of intervals between consecutive beats originating from the sinus node [1]. Over the years, several indices for characterizing the dynamic physiological variation of beat-to-beat heart periods have been proposed and used in different scenarios. Some of these indices have become measurement standards [3]. HRV indices can be classified as time-domain, spectral-domain, or non-linear dynamic indices, and their use depends on the target physiological system, condition, or stressor of interest.

The definition of each HRV index is based on the characterization of a time series of consecutive heartbeat periods. This time series is known as the RR time series (when the period between heartbeats is assessed by using an electrocardiogram (ECG) and a proper QRS detector) or as the inter-beat interval (IBI) time series (when assessed by other physiological signals that are triggered by heart contraction such as finger or wrist photoplethysmography (PPG)). Whatever the IBI or the RR time series is employed, the importance of an accurate estimation of each sample of the series has been stressed elsewhere [3], [4]. Accordingly, accurate HRV index determination is often obtained in controlled environments while restraining movements and/or using uncomfortable instrumentation to avoid heartbeat misdetections.

In recent years, with the development of technology, smart wearable devices have been developed rapidly in various fields such as health care and health monitoring [5]. In the health care field, wearable devices as portable electronic medical devices are used to perceive, record, analyze, regulate, and intervene in physiological process to maintain health. Moreover, they can be utilized to treat diseases with the support of various technologies for identification, sensing, connecting, and storing in physical servers or in the cloud a large amount of information that is relevant of the subject treatment. Therefore, wearables can be used as ambulatory systems providing detailed and individual information about health status. Heart rate (HR) is one of the most often measured parameters while monitoring vital signs, especially in most mobile health (m-Health) applications employing wearable devices [6]. HR assessment represents a routine part of any complete medical examination due to the heart’s essential role in an individual’s health. Therefore, HR measurement is becoming a part of the regular people lifestyle assessment. Many electronic devices such as smartwatches, exercise equipment, and smartphones are becoming able to measure this parameter accurately. Although measuring HR in wearables is not as accurate as the classical ECG methods, it has become a very popular tool for consumers. Some recent wrist-worn wearables, such as the Apple Watch Series 4 to 8, and Samsung Galaxy Watch 4 are monitoring HR with ECG single-lead electrodes, and are approved as medical devices in some countries. However, this technology is still limited as users have to sit with their watch wearing a wristband resting on a flat surface and by putting a finger from the hand opposite to the watch for 30 s to close the circuit. In recent years, the demand for using PPG sensors to monitor HR has increased due to its simple function, high flexibility, and portability [7]. Despite PPG-based methods are more user-friendly and convenient, ECG-based methods are more precise. One of the most challenging problems with wearables is that they are vulnerable to motion artifacts. In recent years, signal processing techniques such as machine learning approaches, have been successful to reduce the impact of motion artifacts and estimate HR properly [8], [9]. HR estimation from artifact-induced signals has been studied using different techniques such as Fast Fourier Transform (FFT), Adaptive Filtering, Independent Component Analysis (ICA), frequency-domain ICA, Empirical Mode Decomposition (EMD), wavelet denoising methods, spectral subtraction, and Kalman Filtering[10]. After applying these techniques, most wearables provide estimates of heart rate (but no direct assessment of beat-to-beat changes in heart rate) that update at intervals that depend on the design of the device. Each of the reported values of heart rate is the output of an (often not-disclosed) algorithm that summarizes, probably by smoothing, the RR/IBI time series for a certain number of consecutive beats. This generally unknown algorithm acts as a filter that reduces the impact of misdetections while providing meaningful heart rate values.

Owing to the ubiquity of wearables and their inability to directly estimate the RR or IBI time series, it is interesting to check whether some of the short-term HRV indices obtained from the RR or IBI time series can be estimated from the smoothed HR time series provided by wearables. This work starts with the estimation of two of the most commonly employed short-term HRV indices: the standard deviation of the RR/IBI time series (known as the SDNN) and the standard deviation of the differentiated RR/IBI time series (generally referred to as the RMSSD). Both indices were among the recommended short-term time-domain HRV indices. While SDNN reflects all the cyclic components responsible for variability during the recording period, RMSSD estimates high-frequency variations in the heart rate [3]. Because SDNN and RMSSD are directly computed from the time series of intervals between consecutive heartbeats, the prediction of both indices employs the reciprocal of the smoothed HR time series, which we refer to as the smoothed heart period time series (sHP).

Hence, the aim of this study is to test the feasibility of estimating the classical HRV short-term SDNN and RMSSD indices from the sHP time series, where each point of the sHP is obtained by smoothing the RR or IBI intervals during a certain interval. The presented methodology can be easily adapted to any sHP measuring system provided that the algorithm to compute the sHP from the IBI or RR time series is known. Moreover, the methodology can be easily expanded to other HRV indices such as spectral indices.

## Materials and methods

In this work, we attempt to estimate the SDNN and RMSSD indices that quantify the original RR/IBI intervals from the sHP time series. The estimation of SDNN and RMSSD uses the features of the corresponding sHP time series feeding an artificial neural network (ANN), whose characteristics depend on the smoothing algorithm applied to the original RR/IBI intervals. The recording duration for each time series was approximately 5 min. Fig 1 shows the methodology that we followed and described in this section.

**Fig 1.**
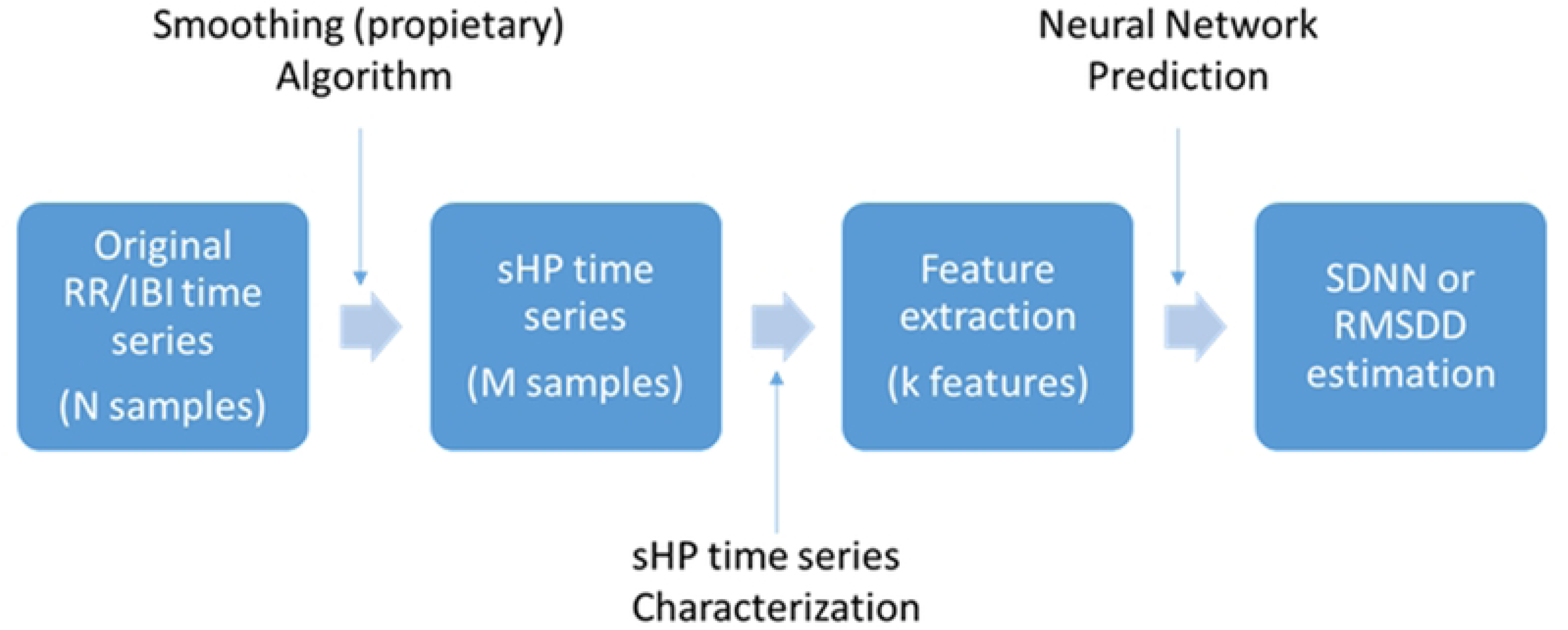
General structure of the proposed methodology.

First, most wearables that measure HR internally measure the heart period (HP) by smoothing the inter-beat intervals of the subject under measurement using an internal algorithm. As shown in Fig 1, during an observational time (5 min in this work), a total of N inter-beat intervals were detected and processed, and an sHP time series with M samples (very often M<N) was obtained. Because the processing procedure is not generally disclosed in commercial devices, this work presents the results for some tentative smoothing algorithms. Most devices record the evolution of the sHP over time. These time series are characterized in this work using simple statistical indices to obtain a total of k features for each sHP time series during the observational time. These features are fed to an ANN with k inputs and one output that provides an estimation for either the SDNN or RMSSD of the original RR/IBI time series. The ANN depends on the smoothing algorithm, selected features, and intended index to be estimated.

To test the accuracy of the SDNN and RMSSD estimates that can be obtained from actual recordings, we used the following methodology:

1.- A large number of RR time series with durations longer than 5 min were obtained from available free ECG databases or annotations. These databases and the RR time-series procurement are described in Section 2.1.
2.- The selected time series was split into non-overlapping sections with durations between 4.5 and 5 minutes. Avoiding overlap among split sections guarantees that the training, test, and validation sets for ANN fitting contain information corresponding to different feature realizations. Almost each employed RR time series section had a duration very close to 5 min; however, if the last non-overlapping section associated with the recording of a subject lasted more than 4.5 minutes, it was also included in the analysis. For each section, the sHP time series is computed using the proposed smoothing algorithm. The splitting and smoothing algorithm proposals are presented in Section 2.2.
3.- A total of 16 features have been employed to characterize each sHP time series. The features are described in section 2.3
4. For each smoothing algorithm and target index (SDNN or RMSSD), an ANN was trained and tested. The structure of the ANN, learning procedure, and validation and testing stages are presented in section 2.4 as well as the statistics employed to quantify the differences between the estimated indices and the indices obtained from the original RR/IBI time series.

### Databases description and RR time series procurement

Three databases available at the Physionet site [11] were used in this study. All three databases contained at least one channel of raw ECG on healthy volunteers, measured for at least 8 min.

The Autonomic Aging database [12] contains at least one channel of ECG measured at rest during an average of 19 min (ranging from 8 min to 45 min) of 1121 healthy volunteers with ages ranging from 18 to 92 years. The ECG signal was sampled at 1 kHz. Some of the database recordings had two ECG channels. For detection, the first ECG channel (ECG1) was employed; however, the second channel was used when the quality of ECG1 was qualified as very poor by visual inspection and the second channel offered a significantly better quality. For all volunteers, the RR time series was obtained using the QRS detector included in the Kubios HRV Premium (3.5.0), which interpolates the input signal to obtain an equivalent sampling frequency of 2 kHz [13]. After QRS detection, an automatic artifact correction utility embedded in the same software [14] was employed to obtain the final RR time series. Noise segments detected by software using a medium automatic detector were visually inspected. If the signal was considered noisy because of the presence of short-duration arrhythmia, the segment was corrected using an automatic artifact correction algorithm. In case of noise caused by very poor ECG quality, manual correction of the beats was attempted. Only recordings with considerably poor quality or persistent arrhythmia were excluded from the analysis. These rejected recordings correspond to subjects 0167, 0186, 0244, 0299, 0300, 0304, 0321, 0332, 0365, 0373, 0400, 0428, 0554, 0581, 0604, 0634, 0649, 0653, 0686, 0753, 0767, 0895, 0935 and 1011. Some short segments of the detected RR time series were deemed as noise by the Kubios software and were assigned a Not a Number value in the corresponding output file. These segments were cropped prior to analysis. Finally, 1097 RR time series were included in the study.

The Fantasia Database [15] contains ECG recordings of 40 healthy subjects measured while watching Disney’s Fantasia movie. The ECG signal was sampled at 250 Hz. The ages of the subjects ranged from 21 to 85 years. RR time series detection follows the same methodology as in the Autonomic Aging database; therefore, the signal is interpolated to have an equivalent sampling frequency of 2 kHz. Recording f2o08 was rejected because of the persistence of arrhythmia, while recording f2y10 was rejected because the signal was lost during some long segments of the recording. In total, 38 RR time series were included in this study.

The Normal Sinus Rhythm RR Interval Database [11] contains beat annotation files for 54 long-term ambulatory ECG recordings of subjects with normal sinus rhythm while performing their normal activities. The original ECG recordings from which annotations were obtained had a sampling frequency of 128 Hz. Hence, the RR time series of this database has a lower time resolution than that of the other databases. The ages of the subjects ranged from 28 to 76 years. After reading the annotations with software available on the PhysioNet web (using the “rdann” function and Matlab© [16]), the raw RR time series were obtained by differentiating the location of the annotations. Then, the corrected RR time series was obtained using the Kubios HRV Premium (3.5.0) software using the automatic correction algorithm. The automatic noise detection was set to a medium level, and zones that were classified as noise were cropped and not considered for analysis. The nsr024 recording was rejected for the analysis because it showed too many ectopic beats. Accordingly, 53 RR time series were included in this study.

The RR time series of the three databases and their corresponding time vectors from the beginning of each recording are available in this public repository.

### Smoothed Heart Period time series definitions

Each recording in the repository consisted of two vectors: a vector ***t*** containing timestamps and their corresponding ***RR*** intervals. Each timestamp was obtained as the arithmetic mean of two consecutive QRS locations, and the corresponding RR interval was obtained as the difference between them. A general smoothing algorithm looks for the samples in the ***RR*** time series that start at timestamp *t_min_* and end at timestamp *t_max_* and computes a number reflecting the central tendency of the selected RR samples. Updating the values of *t_min_* and *t_max_* produces different central-tendency numbers. Therefore, a smoothed heart period time series (***sHP***) was obtained by changing the values of the start and finish times.

Here, for each RR time series in the repository, the ***sHP*** time series is generated using an iterative procedure with initial values of *t_min_*(0) = 0 s and *t_max_*(0) = *T* and these values are updated for each iteration as *t_min_*(*i*+1) = *t_min_*(*i*)+*Δt* and *t_max_*(*i*+1) = *t_max_*(*i*)+*Δt*. In each iteration, the central tendency of the samples in the ***RR*** time series with associated timestamps between *t_min_* and *t_max_* is computed, and the corresponding measurement timestamp is determined as *t_sHP_*(*i*)=(*t_min_*(*i*)+ *t_max_*(*i*))/2. In this study, to assess the influence of the smoothing procedure, two combinations of *T* and *Δt* (that will referred to as the sampling strategies) were employed:

SS1 or sampling strategy #1: *T*= 10 s, *Δt* = 1 s

SS2 or sampling strategy #2: *T*= 30 s, *Δt* = 5 s

We also employed four central tendency measures to characterize the selected RR time intervals to define the ***sHP*** time series:

CTM1 or central tendency measure #1: The arithmetic mean of the RR time intervals starting at *t_min_* and ending at *t_max_*

CTM2 or central tendency measure #2: The median of the RR time intervals starting at *t_min_* and ending at *t_max_*. This is a robust measure against outliers in the RR time series.

CTM3 or central tendency measure #3: This central tendency mimics when employing averaged heart rate time series from commercial devices that are normally quantified as integers in beats per minute (bpm). If the RR time series is in milliseconds,

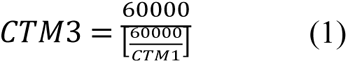

CTM4 or central tendency measure #4: As in the case of CTM3, but using the median instead of the arithmetic mean to perform the rounding:

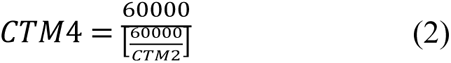

Fig 2 shows an example of how the ***sHP*** time series was obtained using SS1 and CTM1. Note that, although the ***RR*** time series is an unevenly sampled time series, the ***sHP*** time series is evenly sampled when using the proposed sampling strategies.

**Fig 2.**
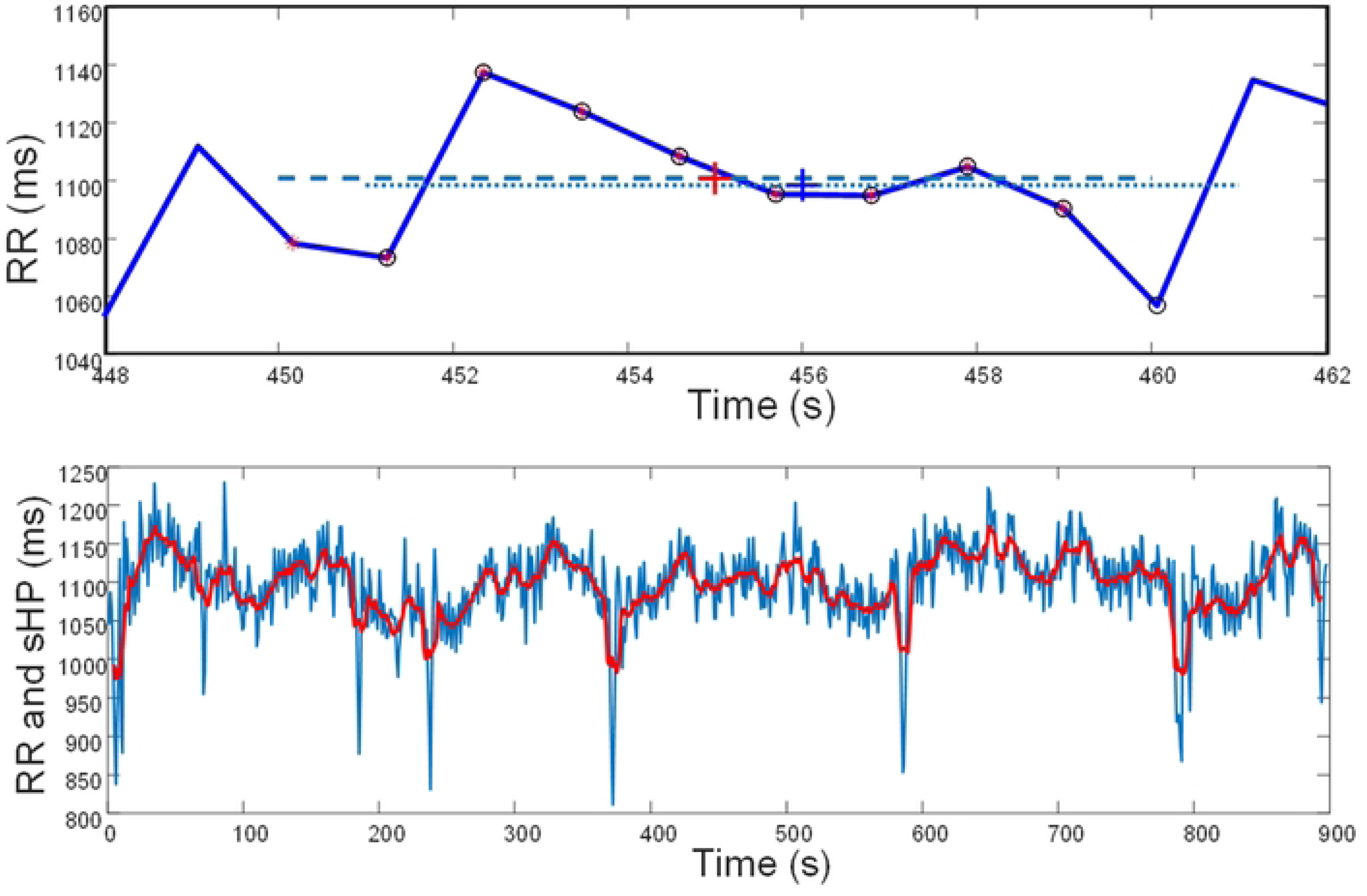
Example of computation of *sHP* using the sampling strategy #1 (window length *T*= 10 s and sliding step *Δt* =1 s) and central tendency measure #1 (arithmetic mean) for the subject 1107 of the Autonomic Aging database. The upper panel shows the details of the computation of the central tendency in a short segment of the recording. The red asterisks show the *RR* time intervals used for the computation of the *sHP* for the window starting at 450 s and ending at 460s while the blue circles show the *RR* time intervals used for the next iteration (starting at 451 s and ending at 461 s). The dashed and dotted lines show the time intervals for smoothing and the corresponding arithmetic means. The red and blue crosses reflect the two arithmetic means located at the center value of the measurement interval (455 s for the first interval, 456 s for the next interval). The lower panel shows the *RR* time series (blue) and the corresponding *sHP* (red) after iteratively applying the sampling strategy #1 and computing the central tendency measure #1 through the whole recording.

### Target computation, *sHP* segmentation and feature extraction

SDNN and RMSSD should be computed for approximately the same recording length. In the short-term HRV analysis, this was approximately 5 min. Nevertheless, the ***RR*** time series in the repository ranged from 8 min to more than 24 h. Hence, it is necessary to partition the ***RR*** time series obtaining an approximately 5 minutes long time series and compute the short-term HRV time indices from them. SDNN and RMSSD were the target indices in this study. In parallel, the segments of the ***sHP*** time series that originate from the partitioned ***RR*** time series must be identified. Selected features from these ***sHP*** segments will feed the designed machine-learning algorithms to estimate the corresponding target HRV time indices. This procedure was performed as follows.

1. Initially, an observational window is located between *t_start_*=0 s and *t_end_*= 300 s.
2. The samples of the ***RR*** time series that have corresponding ***t*** timestamps inside the interval [*t_start_*, *t_end_*] are used to compute SDNN and RMSSD, as explained in [3].
3. An ***sHP_s_***time series is cropped from the ***sHP*** time series by identifying the samples that satisfy their corresponding ***t_sHP_*** timestamps and are included inside the interval [*t_start_*,*t_end_*].
4. The selected features that will be described next are extracted from the ***sHP_s_***. Hence, for each SDNN or RMSSD index, a set of features characterizing the ***sHP_s_***is available.
5. The observation window was displaced by 300 s. If *i* represents the number of iterations, *t_start_*(*i*+1)=*t_start_*(*i*)+300 s and *t_end_*(*i*+1)= *t_end_*(*i*)+300 s
6. While *t_end_*(*i*+1) is lower than the total recording time (maximum of the ***t*** time series), Steps 2, 3, 4, and 5 are repeated.
7. If *t_start_*(*i*+1) is lower than the total recording time, then *t_end_*(*i*+1) is not

a. If *t_end_*(*i*+1)- *t_start_*(*i*+1)≥270 s, repeat one last time the steps 2,3,4 and 5, and the procedure stops.
b. If *t_end_*(*i*+1)- *t_star_*_t_(*i*+1)<270 s, the procedure stops.

Fig 3 shows an example of the procedure using the same recording as the lower panel of Fig 2. In the second iteration, time series with timestamps between 300 and 600 s were selected. The section of the ***RR*** time series is employed to compute the SDNN and RMSSD, whereas the section of the ***sHP*** time series is employed for feature extraction.

**Fig 3.**
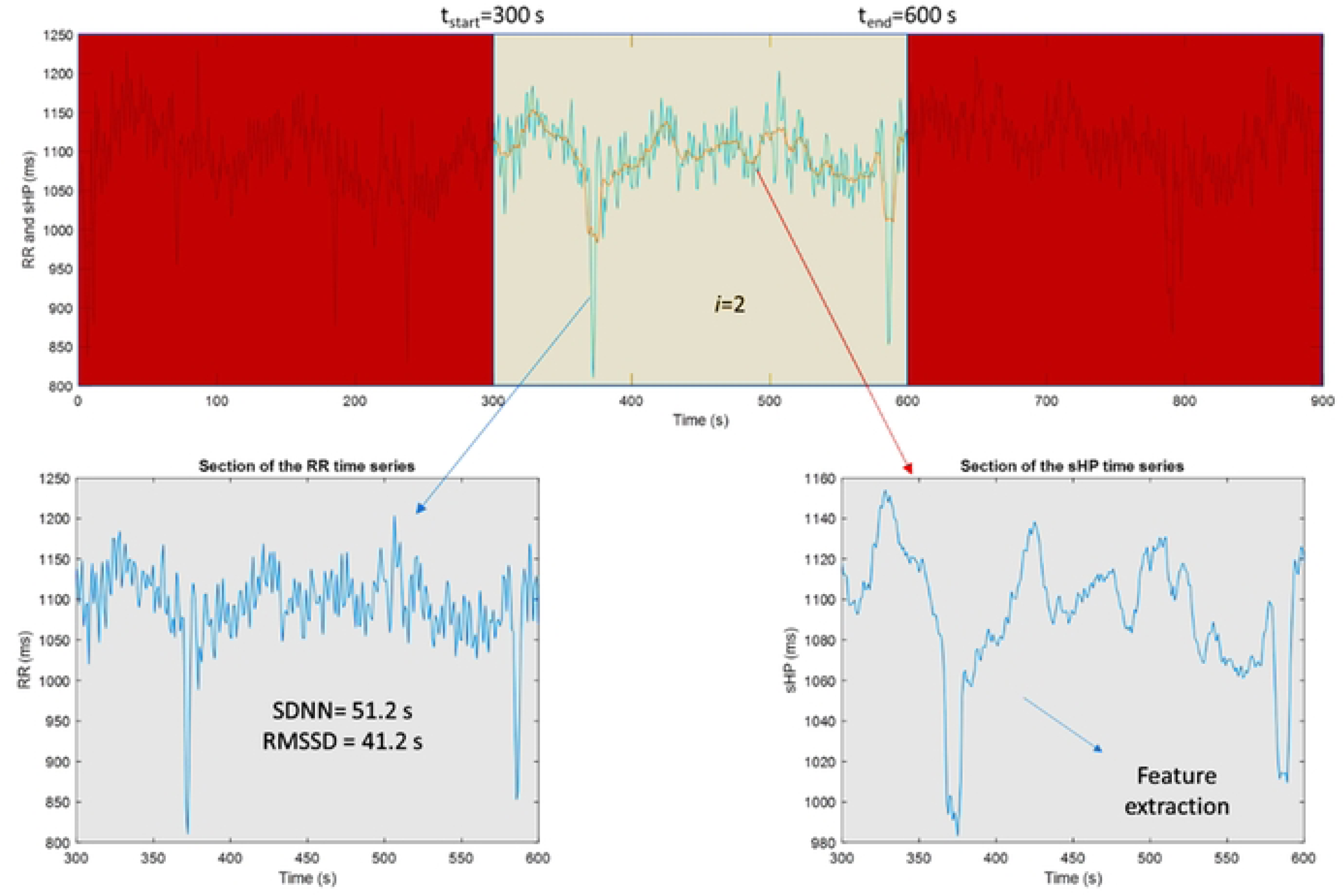
Example of RR index target computation and *sHP* segmentation for the subject 1107 of the Autonomic Aging database. The upper panel shows the observational window for the second iteration (*i*=2) starting at 300 s and ending at 600 s as well as the ***RR*** and ***sHP*** time series. The left lower panel shows the section of the RR time series that correspond to the observational window as well as the values of the target indices (SDNN and RMSSD). The right lower panel shows the section of the ***sHP*** that will be further quantified using some selected features.

Prior to feature extraction, three auxiliary time series are derived from the ***sHP_s_***:

a. The differentiation of the ***sHP_s_*** defined as:

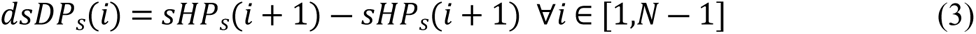

b. The second order differentiation of the ***sHP_s_*** defined as:

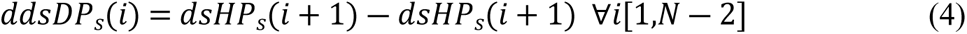

c. The third order differentiation of the ***sHP_s_*** defined as:

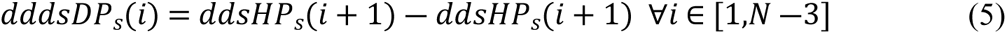

d. The cumulated sum of the ***sHP_s_***after mean removal defined as

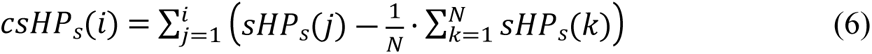

where *N* is the number of samples in ***sHP_s_***. In this study, we obtained 16 features corresponding to the mean value of the ***sHP_s_*** and the sample standard deviation, skewness, and kurtosis of ***sHP_s_***, ***dsHP_s_***, ***ddsHP_s_***, ***dddsHP_s_*** and ***csHP_s_***. Fig 4 shows an example of Figs 2 and 3 showing four of the time series (***dddsHP_s_*** is not shown for the sake of simplicity) and the corresponding values of the features.

**FIG 4.**
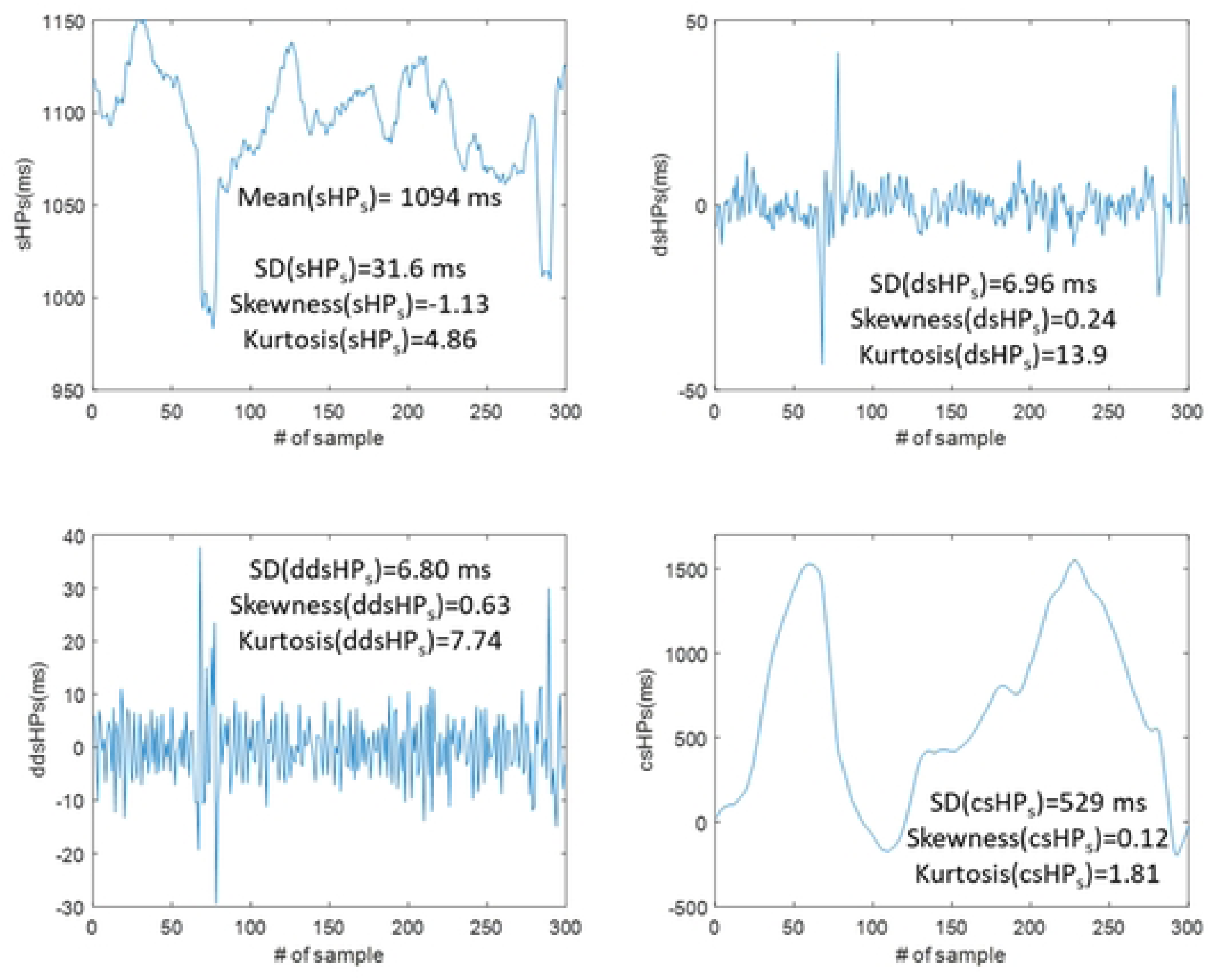
Example of feature extraction for the second segmentation of the *sHP* time series of the subject 1107 of the Autonomic Aging database. The panels show different time series and their corresponding features.

In summary, each ***RR*** time series and ***sHP*** time series were segmented in sections with a duration of approximately 5 min (at least 270 s). Each segment of the ***RR*** time series was used to compute the corresponding SDNN and RMSSD indices. These values will be employed later to train and test a machine learning algorithm. By contrast, each segment of the ***sHP*** defines four auxiliary time series, and three features are obtained from each time series (including the original ***sHP*** segment). These features and the mean value of the ***sHP*** will be the inputs of the machine-learning algorithm, as shown in Fig 1. The functions developed for MATLAB ^©^ that use an arbitrary input ***RR*** time series and their corresponding timestamp time series, generate the ***sHP*** using one of the described sampling strategies and one of the proposed central tendency measures, perform the segmentation, and compute the target indices, which are also available in the public repository. MATLAB files containing the target values and different 16 features for all recordings using the two sampling strategies and four central tendency measures are also available at the repository.

### ANN fitting and testing

In this study, shallow ANNs [17] (with only one hidden neuron layer) were employed to provide estimates of SDNN and RMSSD from the 16 features previously described. The ANNs were trained using the Bayesian regularization backpropagation method [18] to obtain estimates that generalize well. We used shallow instead of deep ANNs to simplify the tailoring of the architecture of the ANNs because the number of hidden neurons is not known a priori. Consequently, several sizes of the hidden neuron layer were tested to choose the size that provides a low error in the estimation while still providing a general solution to the problem. The Deep Learning Toolbox of Matlab^®^ was employed to define, train, validate, test, and evaluate the generalization using the estimation errors of the ANNs.

The size of the hidden neuron layer is a parameter that must be fixed before the start of supervised learning. Because this size may impact overfitting of the model[19], we tested the performance of models with different hidden neuron layer sizes using a subset of subjects, features, and targets from the pool of recordings of the previously described databases. Hence, before starting the learning procedure for any of the ANNs, the features and targets corresponding to approximately half of the subjects in each database were kept for further performance testing. This set of information is referred to as the *keeping* set, while the set employed for the learning of the ANN is referred to as the *learning* set. The MATLAB ^®^ code and the permutation for assigning the subjects to the keeping or learning sets are available <u>at the repository</u>. The same permutation was employed for all the sampling strategies, smoothing algorithms, and target indices (either SDNN or RMSSD); therefore, every ANN in this work learned using the same information. Finally, the learning set batch size was 9317 (obtained from 9317 sections of ***sHP*** time series of durations around 5 min) while the batch size was 9798. The batch size was different for the two sets because the lengths of the recordings were different among the subjects.

For the optimization of the hidden neuron layer size, for every sampling strategy and central tendency measure, we tested hidden neuron layer sizes ranging from 1 to 20. Each model learned using a training set that contained all the batches (features and targets) of approximately 50% of the subjects for each database of the learning set, a validation set that contained batches of approximately 25% of the learning set, and a testing set with the remnants of the learning set. A random permutation allocated each subject of the learning set to the training, validation, or testing sets every time a new model for the ANN was fitted. The learning algorithm used the mean squared error (MSE) between the target and ANN output to fit the model. The hidden neurons made a weighted sum at their inputs and obtained their output using the hyperbolic tangent sigmoid transfer function to accelerate convergence[20]. The code for training, testing, and validating the ANNs using the training set is available <u>in the repository</u>.

Errors in the estimation may not be normally distributed (in fact, visual inspection of targets and estimations in some cases show that they correspond to heavy-tailed distributions). Therefore, for the choice of the hidden neuron layer size, the interquartile range of the errors after fitting the model was employed as a figure of merit. The interquartile range when applying the model to the learning set (IQR_learning_) will be considered as a quantifier of the goodness of fit, and when applying the model to the keeping set (IQR_keeping_) will be considered as a quantifier of the generalizability of the model. Moreover, the interquartile range of the errors when applying the model to the pooling of both sets (IQR_all subjects_) will be considered to optimize the hidden neuron layer size.

For each hidden neuron layer size, we repeated the fitting of the model a number of times equal to the rounding of 300 divided by the hidden neuron layer size, and the IQR_learning_, IQR_keeping_ and IQR_all subjects_ for the realizations, and kept the model with the lowest IQR_all subjects_. We determined the final size of the hidden neuron layer by inspecting the evolution of the IQR_all subjects_ with increasing sizes.

After selecting the hidden neuron layer size, the ANN with the best performance (measured once again by IQR_all subjects_) after 100 completely new fittings was selected as the best ANN for the estimation of the target indices. In total, 16 ANN were obtained (also available <u>at the</u> <u>repository</u> and specified as the Deep Learning Toolbox of Matlab® net variables) corresponding to the combinations of the two target indices, four central tendency measures, and two sampling strategies. IQR_learning_, IQR_keeping_ and IQR_all subjects_ for each ANN were obtained.

Because errors in the estimation of the indices (the estimation error is computed as the difference between the target index and its corresponding estimation obtained from the output of the ANN) are not normally distributed (and in some cases, some outliers may be present), the difference between the 97.5 th and 2.5 th percentiles of the estimation error and the median of the absolute value of the estimation error were also computed for each ANN. Finally, the odds that the absolute or relative estimation errors were lower than a certain threshold were computed for a certain range of thresholds, and the mean values of the odds were obtained for each case. The obtained odds curves and mean odds curve values provide a convenient way to compare the impact of the target index, central tendency measure, and sampling strategy on the performance of different ANN. The MATLAB ^®^ code for all these characterizations is also available <u>in the repository</u>.

## Results

Fig 5 shows the evolution of the IQR_all subjects_ against the hidden neuron layer size for the different sampling strategies (SS), central tendency measures (CTM), and target index. The ordinate axes have different scales to observe for each target and SS, which is a reasonable choice for the hidden neuron layer size. First, the IQR_all subjects_ were lower when estimating SDNN from a device using the SS with a lower smoothing of the data (#1). The worst case occurs when estimating the RMSSD with a large smoothing of data (SS #2). The results were best when using the arithmetic mean for smoothing the data (CTM #1) and worst when using the rounded median (CTM #4). Fig 5 also shows that the IQR_all subjects_ for a hidden neuron layer size of 10 are comparable to those obtained for larger sizes; therefore, it is not necessary to use an ANN with a larger number of neurons. Hence, the remaining results apply to a shallow ANN with 10 neurons in the hidden neuron layer (in this case, selected from the best performance ANN in 100 fittings for each type of target, CTM, and SS).

**Fig 5.**
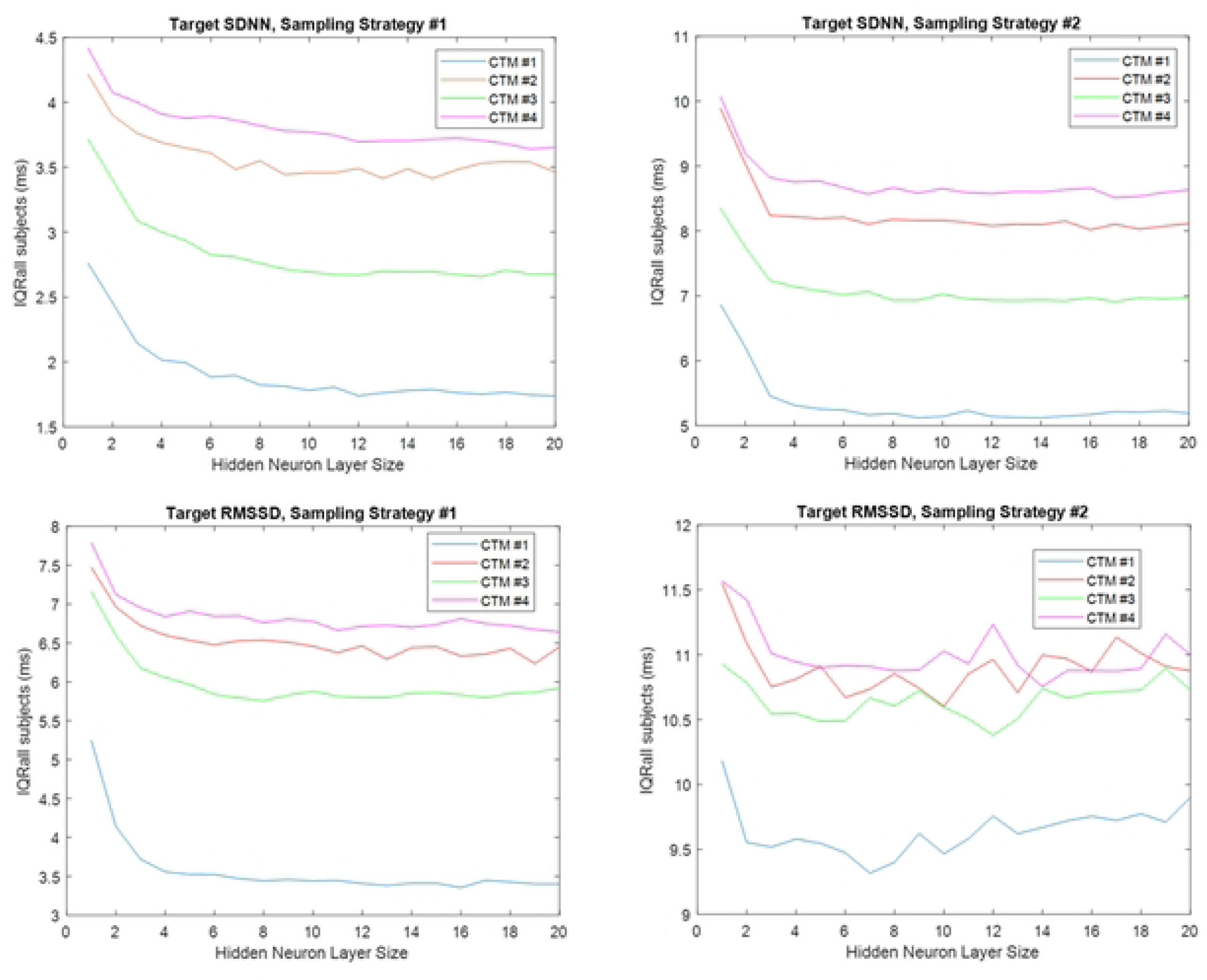
Change of IQR_all subjects_ with the hidden neuron layer size for the two targets (SDNN for the upper panels and RMSSD for the lower panels) and the two sampling strategies (smoothing window of 10 seconds with an update each second for the left panels and smoothing window of 30 seconds with an update every 5 seconds for the right panels) using the 4 analyzed central tendency measures (CTM).

Table 1 lists the IQR_learning_, IQR_keeping_ and IQR_all subjects_ for the different optimized ANN. These results mirror those presented in Fig 5 for a hidden neuron-layer size of 10. Fig 6 shows plots of the value of the target HRV index against the estimation error (difference between this index and the estimated index) to show the agreement of the estimation provided by the ANN. The best and worst cases for each index are provided. The difference in levels of agreement (dLoA) estimated as the difference between the 97.5% and 2.5% percentiles of the estimation error, as well as the median of the absolute value of the estimation error (MAE) for all CTM and SS combinations, are shown in Table 2. The results in Fig 6 and Table 2 use the pooled data of the learning and keeping sets (19115 different targets).

**Fig 6.**
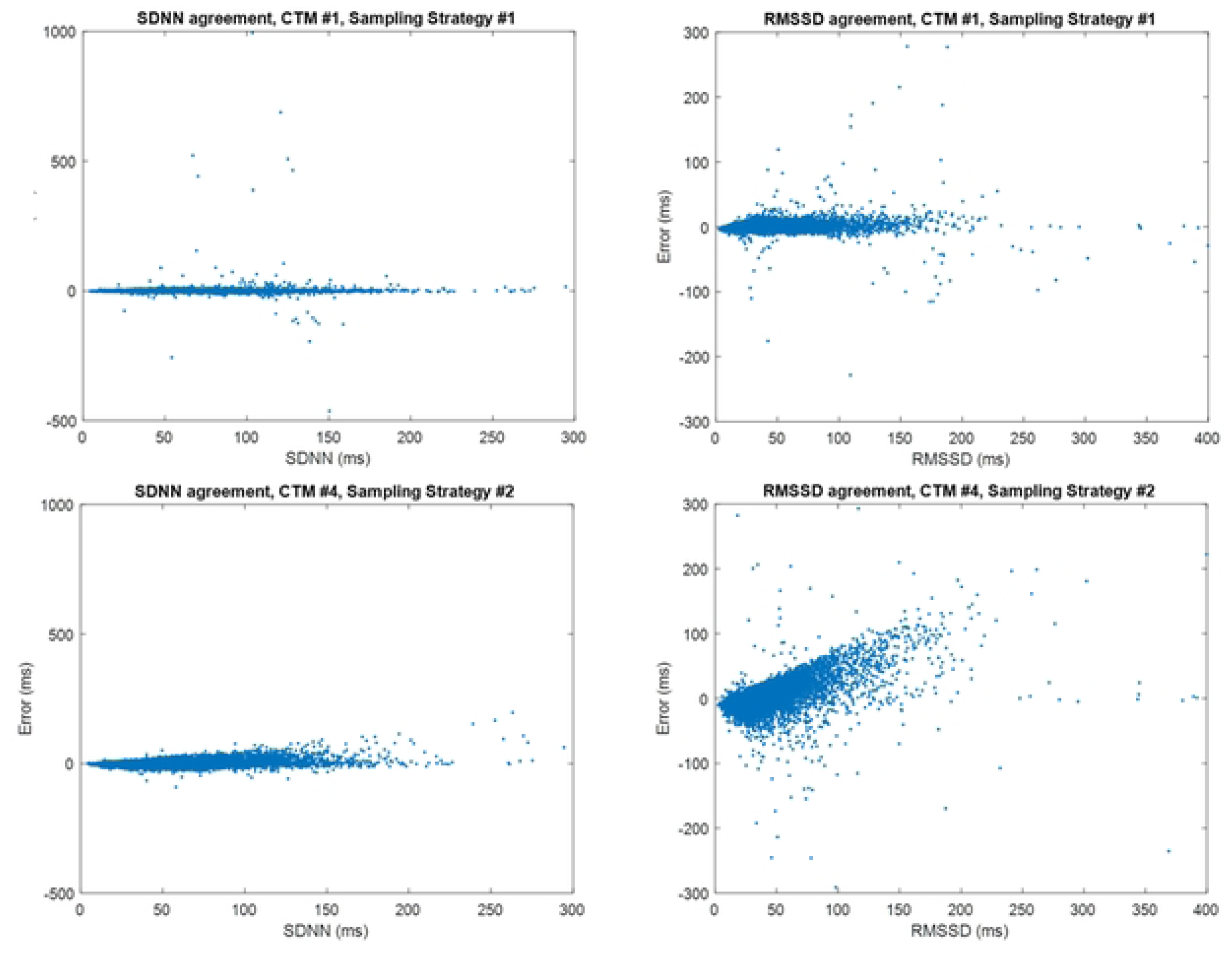
Some agreement plots for the SDNN and RMSSD indices using the best performance ANN and the pooled data of the learning and keeping sets. The upper panels show the estimation error for the best case (CTM #1 and SS #1) while the lower panels show the worst case (CTM #4 and SS #2).

**Table 1.** Interquartile range of the estimation errors for each target index, central tendency measure (CTM) and sampling strategy (SS) assessed using only the learning set (IQR_learning_), the keeping set (IQR_keeping_) or pooling both sets (IQR_all subjects_)

| Target index: SDNN |  |  |  |  |  |  |  |  |
| --- | --- | --- | --- | --- | --- | --- | --- | --- |
| CTM | #1 | #2 | #3 | #4 | #1 | #2 | #3 | #4 |
| SS | #1 | #1 | #1 | #1 | #2 | #2 | #2 | #2 |
| $IQR_{\text{learning}}$ (ms) | 1.77 | 3.29 | 2.68 | 3.56 | 4.94 | 7.77 | 6.72 | 8.23 |
| $IQR_{\text{keeping}}$ (ms) | 1.91 | 3.62 | 2.77 | 4.03 | 5.40 | 8.30 | 7.18 | 9.03 |
| $IQR_{\text{all subjects}}$ (ms) | 1.84 | 3.46 | 2.73 | 3.79 | 5.17 | 8.05 | 6.94 | 8.59 |
| Target index: RMSSD |  |  |  |  |  |  |  |  |
| CTM | #1 | #2 | #3 | #4 | #1 | #2 | #3 | #4 |
| SS | #1 | #1 | #1 | #1 | #2 | #2 | #2 | #2 |
| $IQR_{\text{learning}}$ (ms) | 3.26 | 6.10 | 5.71 | 6.31 | 9.05 | 9.98 | 9.88 | 10.2 |
| $IQR_{\text{keeping}}$ (ms) | 3.66 | 6.90 | 5.92 | 7.04 | 10.2 | 11.6 | 11.3 | 11.7 |
| $IQR_{\text{all subjects}}$ (ms) | 3.47 | 6.48 | 5.82 | 6.65 | 9.60 | 10.8 | 10.6 | 11.0 |

**Table 2.** Difference in the levels of agreement (dLoA) and median absolute error (MAE)

| Target index: SDNN |  |  |  |  |  |  |  |  |
| --- | --- | --- | --- | --- | --- | --- | --- | --- |
| CTM | #1 | #2 | #3 | #4 | #1 | #2 | #3 | #4 |
| SS | #1 | #1 | #1 | #1 | #2 | #2 | #2 | #2 |
| dLoA (ms) | 9.88 | 18.2 | 12.5 | 18.4 | 30.6 | 35.5 | 36.2 | 37.1 |
| MAE (ms) | 0.93 | 1.73 | 1.36 | 1.92 | 2.66 | 4.00 | 3.55 | 4.28 |
| Target index: RMSSD |  |  |  |  |  |  |  |  |
| CTM | #1 | #2 | #3 | #4 | #1 | #2 | #3 | #4 |
| SS | #1 | #1 | #1 | #1 | #2 | #2 | #2 | #2 |
| dLoA (ms) | 15.8 | 31.8 | 25.6 | 33.9 | 66.2 | 68.1 | 72.1 | 69.2 |
| MAE (ms) | 1.72 | 3.39 | 2.92 | 3.41 | 5.15 | 5.54 | 5.82 | 5.73 |

dLoA is estimated as the difference between the 97.5 and 2.5 percentiles of the estimation error and median absolute error (MAE) is estimated as the median of the absolute value of the estimation error. Results are disclosed for all the combinations of the central tendency measure (CTM) and sampling strategies (SS) for the two analyzed targets (SDNN and RMSSD).

As seen in Fig 6, sometimes the estimation provided by the ANN shows poor agreement with the target value even for the best-case scenario (CTM #1 and SS #1). Nevertheless, the agreement is better than that suggested by the plots, as shown in Table 2. For CTM #1 and SS #1, when estimating SDNN, the estimation error is lower than 9.88 ms in 95% of the cases and lower than 0.93 ms for half the cases. Using the same CTM and SS, when estimating the RMSDD, the estimation error is lower than 15.8 ms for 95% of the cases and lower than 1.72 ms for half of the cases. Because of the presence of outliers and, to better characterize the agreement between estimates and target indices, the odds of having an absolute value of the estimation error lower than a fixed threshold and the odds of having an absolute value of the relative estimation error (normalized by the target value of the index) lower than a fixed percentage were computed for each evaluated target index, CTM, and SS. For the absolute value of the estimation errors thresholds from 0 to 100 ms have been considered in steps of 0.01 ms to obtain the odds curve. For the absolute value of the relative estimation error, thresholds from 0% to 100% were employed in steps of 0.01%. Odds curves were computed separately for the learning and keeping sets. Fig 7 shows the results for the best and worst cases, as shown in Fig 6. Table 3 quantifies the odds curves for all combinations of the CTM, SS, and target indices using the arithmetic mean of the odds. An ideal estimation with no estimation error would provide a mean of the odds curve equal to one; hence, the lower the mean odds value, the poorer the estimation.

**Fig 7.**
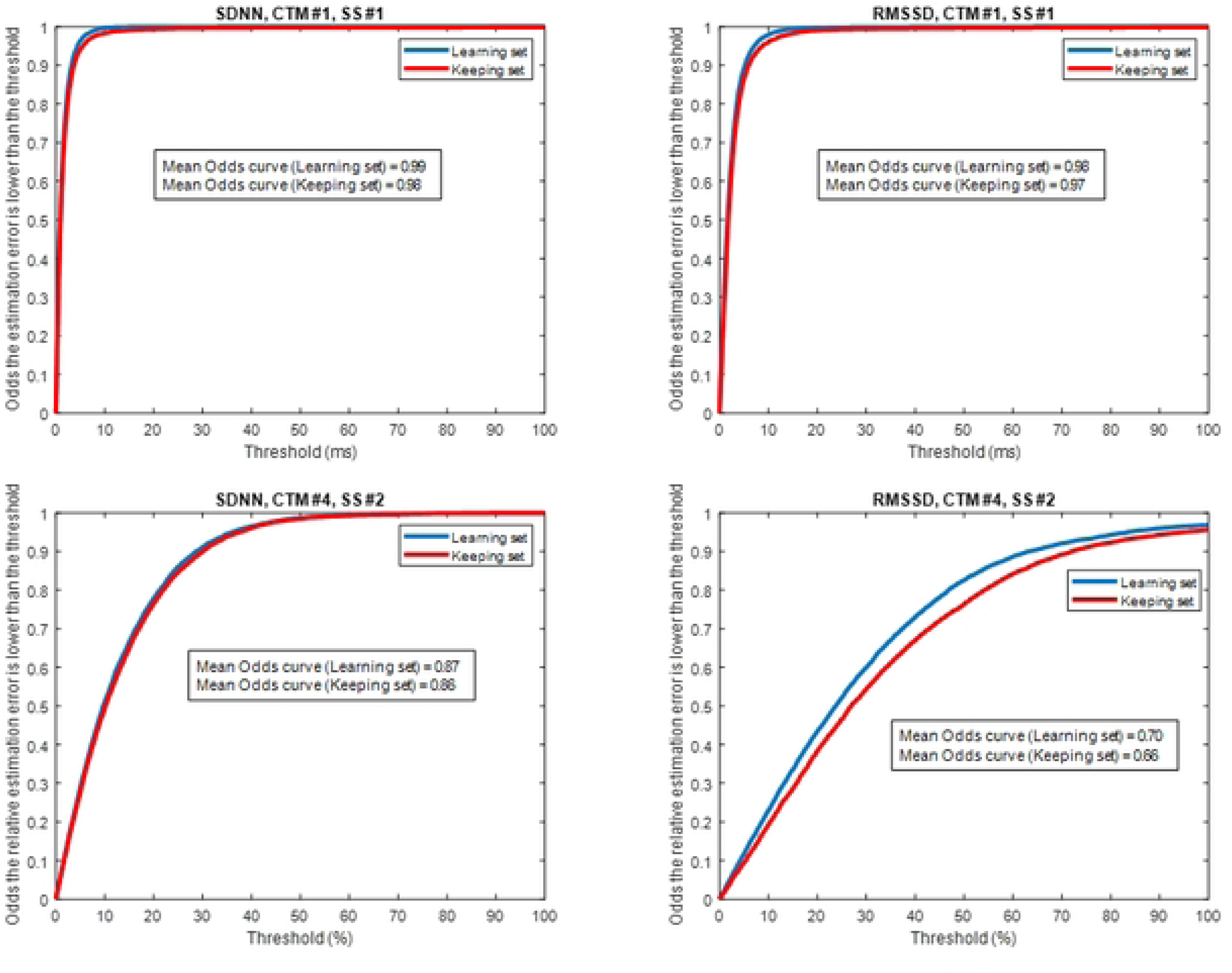
Some Odds curves for the SDNN and RMSSD indices using the best performance ANN reported for the learning and keeping sets. The upper panels show the curves for the best case (CTM #1 and sampling strategy #1) when measuring the absolute estimation error while the lower panels show the worst case (CTM #4 and sampling strategy #2) for the relative estimation error. For example, when using CTM #4 and SS #2 and estimating SDNN, the odds that the relative estimation error is lower than 10% is around 50% for both sets. When using CTM #1 and SS #1 and estimating RMSSD, the odds that the absolute estimation error is lower than 10 ms is around 95% for the keeping set and 97% for the learning set.

**Table 3.** Mean of the Odds curve.

| Mean of the odds curve for target index SDNN |  |  |  |  |  |  |  |  |
| --- | --- | --- | --- | --- | --- | --- | --- | --- |
| CTM | #1 | #2 | #3 | #4 | #1 | #2 | #3 | #4 |
| SS | #1 | #1 | #1 | #1 | #2 | #2 | #2 | #2 |
| LA | 0.99 | 0.97 | 0.98 | 0.97 | 0.96 | 0.94 | 0.95 | 0.94 |
| KA | 0.98 | 0.96 | 0.98 | 0.97 | 0.95 | 0.94 | 0.94 | 0.93 |
| LR | 0.97 | 0.94 | 0.95 | 0.94 | 0.91 | 0.88 | 0.88 | 0.87 |
| KR | 0.96 | 0.93 | 0.95 | 0.93 | 0.91 | 0.87 | 0.88 | 0.86 |
| Mean of the odds curve for target index RMSSD |  |  |  |  |  |  |  |  |
| CTM | #1 | #2 | #3 | #4 | #1 | #2 | #3 | #4 |
| SS | #1 | #1 | #1 | #1 | #2 | #2 | #2 | #2 |
| LA | 0.98 | 0.95 | 0.96 | 0.95 | 0.92 | 0.91 | 0.91 | 0.91 |
| KA | 0.97 | 0.94 | 0.95 | 0.94 | 0.90 | 0.89 | 0.89 | 0.89 |
| LR | 0.91 | 0.83 | 0.85 | 0.82 | 0.73 | 0.71 | 0.70 | 0.70 |
| KR | 0.90 | 0.81 | 0.83 | 0.80 | 0.69 | 0.67 | 0.66 | 0.66 |

This table considers thresholds in the estimation of the absolute error between 0 to 100 ms (A) or in the relative estimation error between 0 to 100% (R) for the different target indices, central tendency measures (CTM) and sampling strategies (SS). Results are reported separately for the learning (L) and Keeping (K) sets (i.e. KA is the mean of the odds curve for the absolute estimation error for the keeping set)

## Discussion

The results show that it is feasible to estimate the SDNN or RMSSD using the features of ***sHP*** time series and shallow ANN. Moreover, they are reasonable: the estimation of RMSSD is worse than that of SDNN because it reflects high-frequency components that are filtered by the smoothing procedure. Furthermore, the larger the smoothing of the data (window length), the larger are the estimation errors for both indices. Nevertheless, the solutions obtained for the estimation of the indices were far from optimal. First, a shallow ANN was not the best choice. As seen in the estimation of RMSSD when using central tendency measure #4 and sampling strategy #2 in Fig 6, the neural network is more prone to provide positive errors with increasing RMSSD values, thus providing lower estimates of the index. In these scenarios, a deep-learning ANN can provide better results. Moreover, while accepting a shallow ANN as a feasible solution, the results always consider the same set of 16 features, which are basic statistical measurements (the first four statistical moments) applied to the ***sHP***, successive differentiation of this time series, and the cumulative sum (after mean removal) of the time series. A different set of features can provide better results, even when using a lower number of features. Further work could be devoted to the search of sets of features that reduce the estimation errors, especially for cases with higher errors, such as when using a large smoothing (i.e., SS #2) for the estimation of RMSSD.

Regarding the employed feature set, some of the features could be irrelevant for building the estimates. To identify irrelevant features, the importance of each feature was tested for each combination of target, CTM, and SS using the increase in the error of the estimation when each of the input features suffers a random permutation [21], creating a mismatch between the feature and its corresponding target. As in the selection of the best neural network, we used the interquartile range of the estimation error by pooling the learning and keeping sets. The importance of feature *j* has been assessed by computing

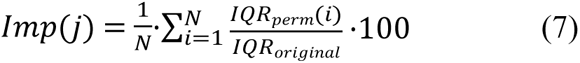

where IQR_perm_(*i*) is the IQR_all subjects_ using the *i*-th random permutation of the input feature *j* (the remaining features are kept in the original order) and IQR_original_ is the IQR_all subjects_ without making any permutation on the input features. *N* is the number of times the feature *j* is permutated. Fig 8 shows bar plots of the feature importance for SDNN and RMSSD when using CTM #1 and #2 and SS #1 and #2 for *N*=100. Table 4 shows, for each target, CTM and SS, which are the most important features, and the list of features that have an importance higher than 300% (*N* = 100). Code for feature importance is also available <u>at the repository</u>. As shown in Fig 8, for a short smoothing window (SS #1) and simple smoothing algorithm (CTM #1), some features are prominent with respect to the others. Nevertheless, as the smoothing window length increases (SS #2) and some rounding and artifact rejection techniques enter the smoothing algorithm (CTM #4), the differences in importance among the features severely decrease. Table 4 shows that for SDNN estimation, the most important feature is, depending on the CTM and SS, the standard deviation of the ***sHP_s_*** time series, or the standard deviation of ***ddsHP_s_***. Other relevant features are the standard deviation of ***dsHP_s_***, standard deviation of ***dddsHP_s_*** and mean value of ***sHP_s_***. For RMSSD estimation, the most important feature is the standard deviation of ***ddsHP_s_*** or the standard deviation of ***dddsHP_s_***. Another important feature is the standard deviation of ***dsHP_s_***. It makes sense that, for the estimation of indices based on the standard deviation of ***RR*** or the standard deviation of the first differentiation of ***RR***, the most important features are the standard deviation of differentiated versions of ***sHP_s_***. Nevertheless, the importance of the features decreased when changing from SS #1 to SS #2. This suggests that the algorithm for estimating the indices for SS #2 is more complex because there is no small set of features that can be considered important, and any of the features can provide useful information to build the estimates. In summary, it seems that the use of the standard deviation of differentiated versions of the ***sHP_s_*** time series probes is useful for the estimation of SDNN and RMSSD for short smoothing periods, where errors in the estimation are generally low (as seen in Tables 1 to 3). Nevertheless, errors in the estimation increased when SS #2 was employed. In these cases, other features can probably improve the estimation of indices.

**Fig 8.**
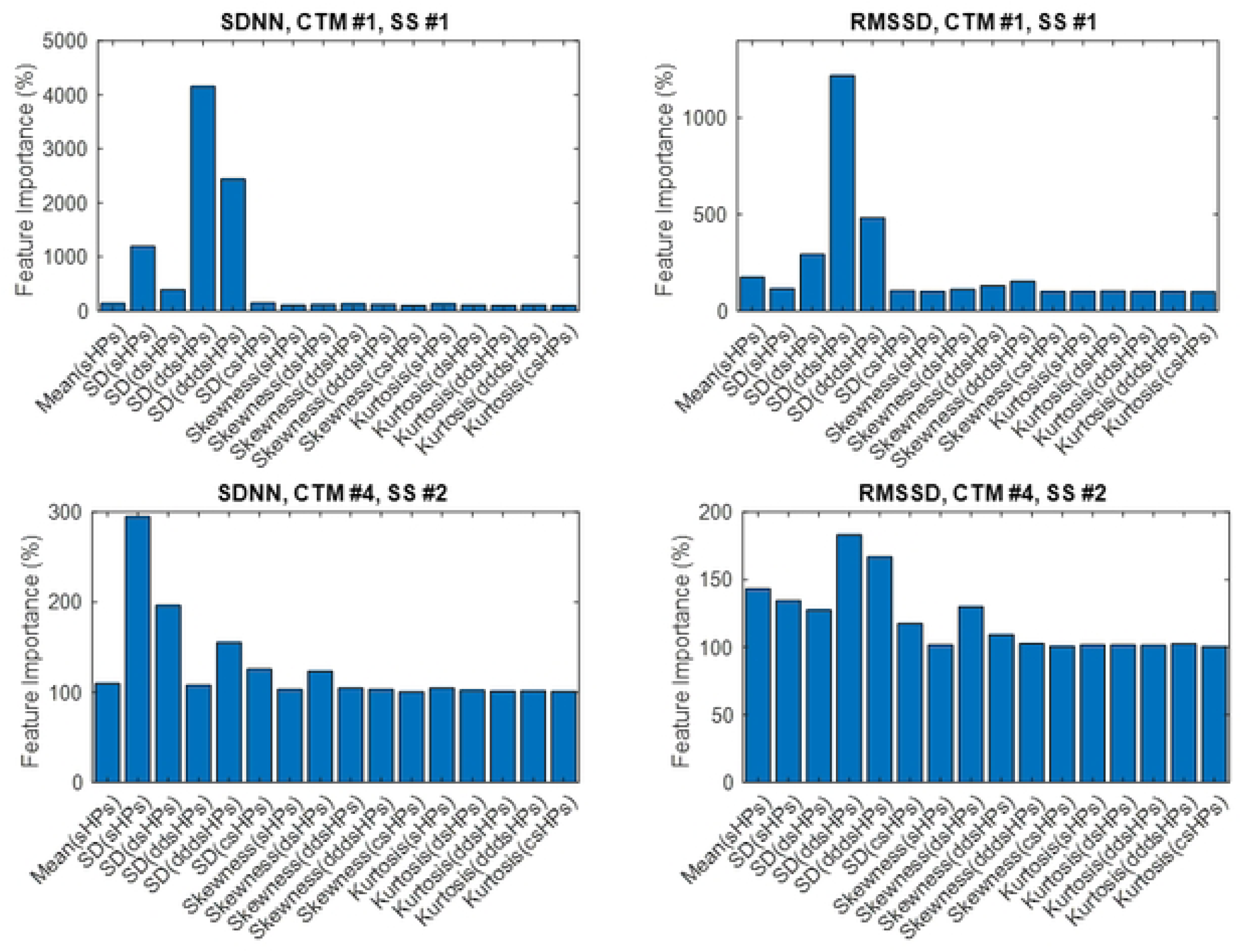
Feature importance for some selected combinations of target, CTM and SS. The two upper panels provide the feature importance when using CTM #1 and SS #1 for SDNN and RMSSD while the two lower panels provide the feature importance for CTM #4 and SS #2 for SDNN and RMSSD.

**TABLE 4.**
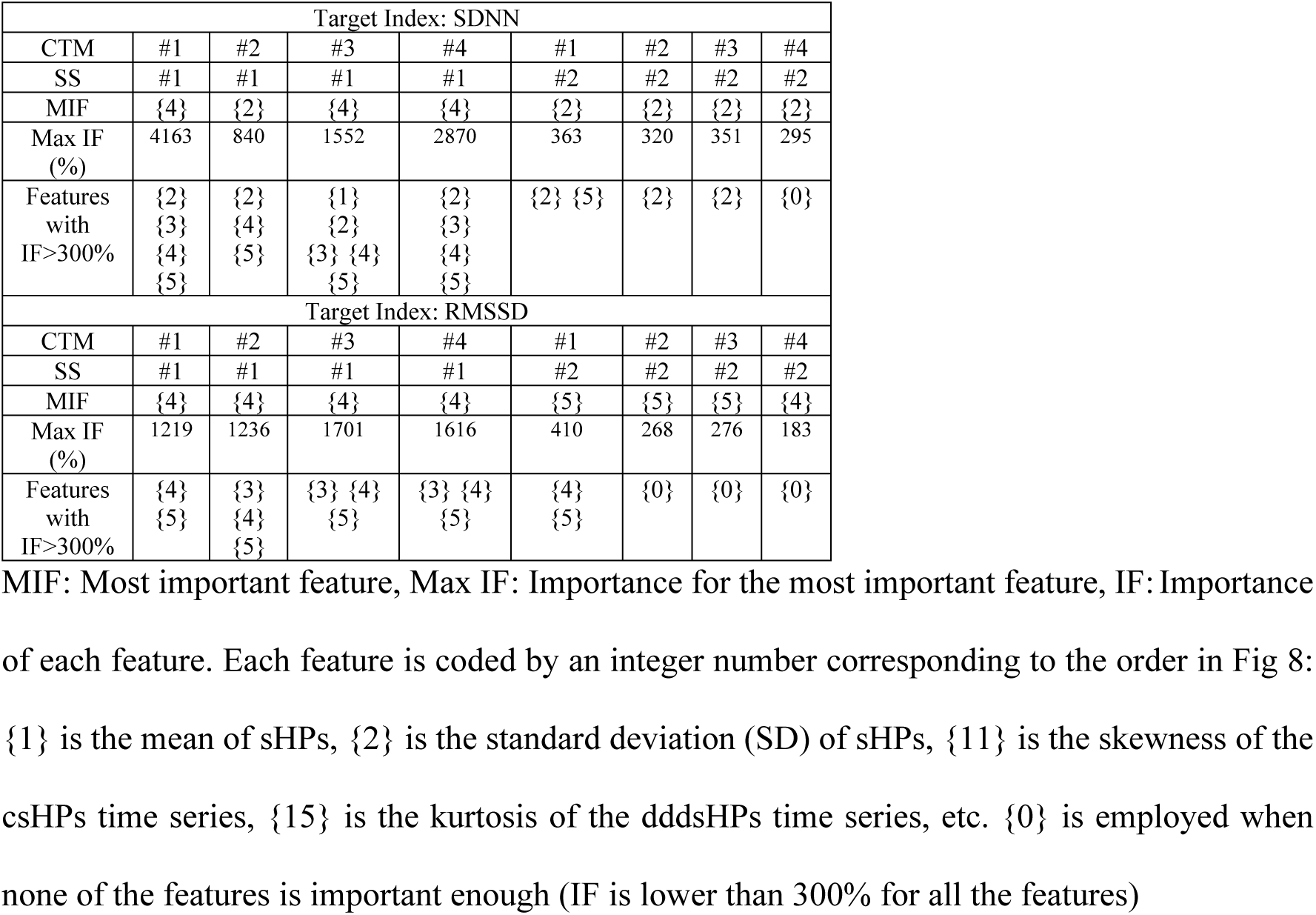
Most relevant results for the feature importance analysis using random permutations.

The main results of this work have dealt with an ANN with a hidden neuron layer size of 10 neurons, which is not a very large number, when using 16 input features. The number of input features was fixed from the beginning of the ANN design. If a reduced set of features is employed (i.e., only using the standard deviation of differentiated versions of ***sHP_s_***) the size of the hidden neuron layer can be changed by either enlarging or stretching. A joint optimization of the number of input features, hidden neuron layer size, and proper selection of features could improve the estimation of the indices, and will be the purpose of future research.

To train the ANN, the Bayesian regularization backpropagation method was used for the sake of generalizability. As seen in Tables 1 and 3 and in Fig 7, the performance of the ANNs is slightly worse in the keeping than in the learning sets. Because the differences in *IQR* or in the mean of the odds curve are small, we can consider that the estimates could generalize well for a completely new set of input features coming from a new heart rate-measuring device. Nevertheless, to obtain good estimations, the ANN must be tailored to the underlying device algorithm for heart rate (or heart period) estimation. In this study, four different smoothers (CTM) and two different smoothing procedures (SS) were used that can be present in some wearable devices. The methodology can be applied to other algorithms if manufacturers disclose them.

This study employed three different public ECG or beat annotation databases to generate the targets and features. The Autonomic aging database and the Fantasia database were measured while the subjects were at rest, while the normal sinus rhythm RR database corresponded to ambulatory measurements. Although the number of different subjects was overwhelmingly larger for the first database, the number of analyzed 5-minute segments was larger for the normal sinus rhythm RR database (14363 segments from a total of 19115 analyzed segments). Hence, we can consider that most of the features employed for the learning and keeping sets correspond to ambulatory measurements. This affects the performance of the ANNs for each database. Table 5 shows the median of the estimation error and the median of the relative estimation error for different CTM, SS, and target indices for the three databases. The median values among databases using the Kruskal-Wallis test showed very significant differences (p<0.001) for all CMT, SS, and target indices, except for the relative estimation error of RMSSD while using CTM #4 and SS #1. This is a foreseeable result, because most of the information provided to the learning algorithm comes from this database. However, worse results correspond to the Autonomic Aging database. This could be attributed to the large number of different subjects in the database and the wide age range. Hence, the training of the ANNs for future development of SDNN and RMSSD estimators should also be performed using a sample of the population with characteristics as close as possible to the subjects the algorithm is intended to be applied.

**TABLE 5.**
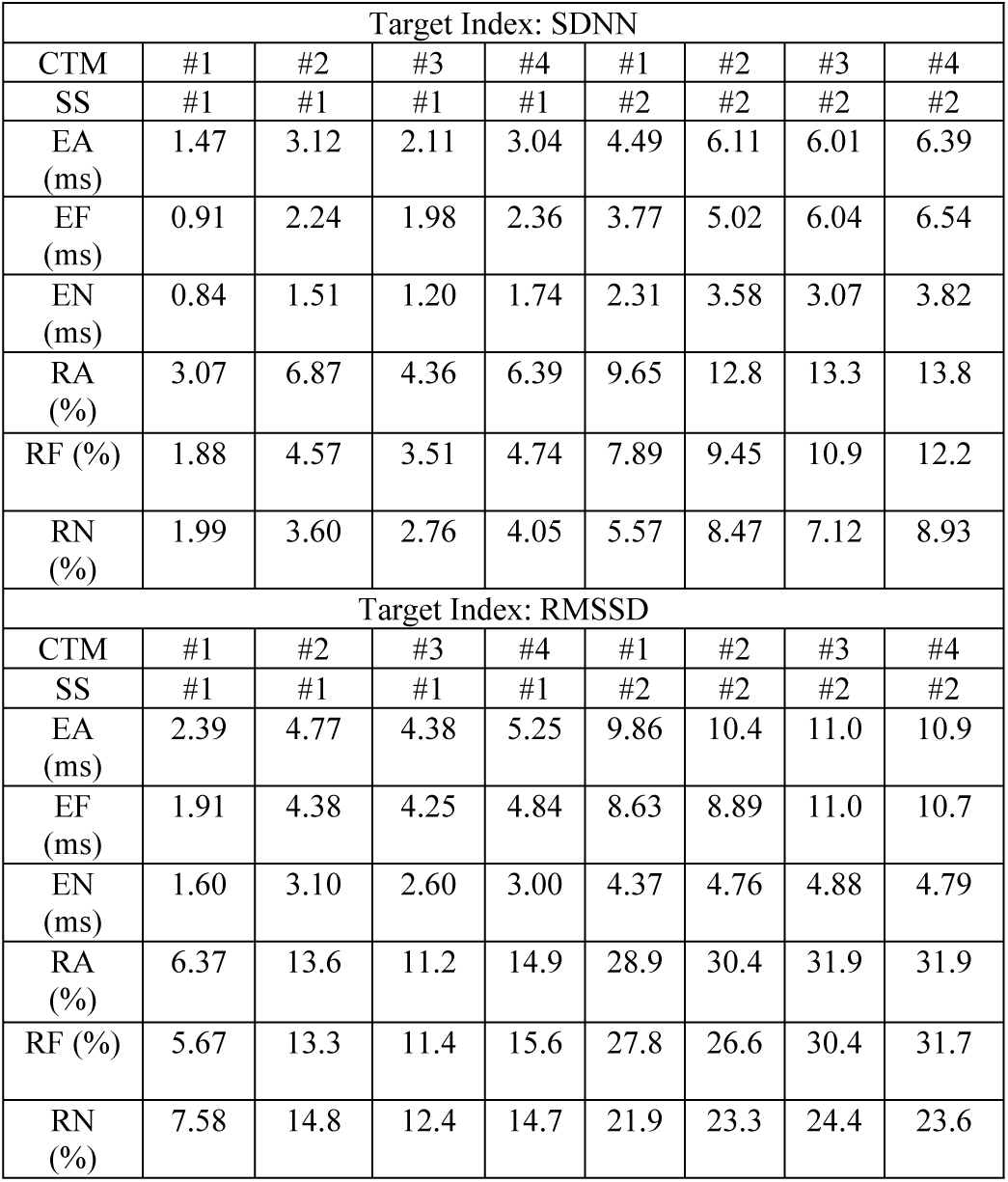
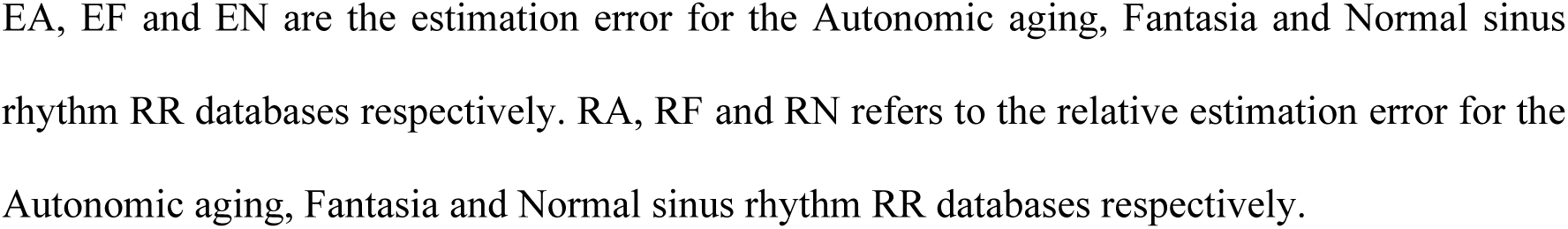
Median values of estimation errors using the best ANNs disclosed by database, target index, CTM and SS.

| Target Index: SDNN |  |  |  |  |  |  |  |  |
| --- | --- | --- | --- | --- | --- | --- | --- | --- |
| CTM | #1 | #2 | #3 | #4 | #1 | #2 | #3 | #4 |
| SS | #1 | #1 | #1 | #1 | #2 | #2 | #2 | #2 |
| EA (ms) | 1.47 | 3.12 | 2.11 | 3.04 | 4.49 | 6.11 | 6.01 | 6.39 |
| EF (ms) | 0.91 | 2.24 | 1.98 | 2.36 | 3.77 | 5.02 | 6.04 | 6.54 |
| EN (ms) | 0.84 | 1.51 | 1.20 | 1.74 | 2.31 | 3.58 | 3.07 | 3.82 |
| RA (%) | 3.07 | 6.87 | 4.36 | 6.39 | 9.65 | 12.8 | 13.3 | 13.8 |
| RF (%) | 1.88 | 4.57 | 3.51 | 4.74 | 7.89 | 9.45 | 10.9 | 12.2 |
| RN (%) | 1.99 | 3.60 | 2.76 | 4.05 | 5.57 | 8.47 | 7.12 | 8.93 |
| Target Index: RMSSD |  |  |  |  |  |  |  |  |
| CTM | #1 | #2 | #3 | #4 | #1 | #2 | #3 | #4 |
| SS | #1 | #1 | #1 | #1 | #2 | #2 | #2 | #2 |
| EA (ms) | 2.39 | 4.77 | 4.38 | 5.25 | 9.86 | 10.4 | 11.0 | 10.9 |
| EF (ms) | 1.91 | 4.38 | 4.25 | 4.84 | 8.63 | 8.89 | 11.0 | 10.7 |
| EN (ms) | 1.60 | 3.10 | 2.60 | 3.00 | 4.37 | 4.76 | 4.88 | 4.79 |
| RA (%) | 6.37 | 13.6 | 11.2 | 14.9 | 28.9 | 30.4 | 31.9 | 31.9 |
| RF (%) | 5.67 | 13.3 | 11.4 | 15.6 | 27.8 | 26.6 | 30.4 | 31.7 |
| RN (%) | 7.58 | 14.8 | 12.4 | 14.7 | 21.9 | 23.3 | 24.4 | 23.6 |

EA, EF and EN are the estimation error for the Autonomic aging, Fantasia and Normal sinus rhythm RR databases respectively. RA, RF and RN refers to the relative estimation error for the Autonomic aging, Fantasia and Normal sinus rhythm RR databases respectively.

All RR time series, features, and targets are available <u>in the repository</u>. Hence, the use of other machine-learning approaches or smoothing algorithms for these data is welcomed.

## Conclusions

This work shows the feasibility of estimating SDNN and RMSSD HRV indices by extracting features from the heart rate (or heart period) time series once a smoothing algorithm has transformed the RR or IBI intervals into a smoother version. The extracted features were fed into a properly fitted ANN to estimate the aforementioned indices. The weights and biases of the ANNs depend on the index to be estimated and the smoothing algorithm. Because the smoothing algorithm made by a particular device is generally not disclosed, this study has proposed eight different procedures based on four different central tendency measures and two different sampling strategies. The results show that RMSSD is harder to estimate than SDNN, and the estimation error increases with smoothing of the RR or IBI time series. Moreover, this depends on the database. Further research on the proposal of new features, their choice, and redesigning of the ANN structure can provide results with lower estimation errors.

## Data Availability

All files and code are available at https://osf.io/f4x89/?view_only=335472277c2241f09f395e763f2b3201

